# Patient and Public involvement in the design of an international clinical trial: Real world experience

**DOI:** 10.1101/2024.09.03.24312062

**Authors:** Gwenda Simons, Helen Jones, Ian Clarke, Firoza Davies, Stacey Grealis, Elspeth Insch, Hameed Kahn, Joanne Lloyd, Al Richards, Hayley Rose, Ruth Williams, Maarten de Wit, Clarissa Woodcock, Leigh Romaniuk, Michelle Bardgett, Arthur G Pratt, Marie Falahee

**Author notes:** Joint senior authors (Arthur G Pratt, Marie Falahee).

## Abstract

**Background:** The value of patient and public involvement (PPI) during the earliest stages of clinical trial development, and prior to the award of substantive funding, is widely recognised. However, it is often under resourced and PPI processes during this phase are rarely reported in detail. Having benefitted from seed funding to develop an international clinical trial proposal, we sought to describe and appraise PPI activities and processes that support pre-award co-development.

**Methods:** A 12-month “accelerator” award facilitated development of a substantive funding application to deliver the Rheumatoid Arthritis Prevention PlatfORm Trial (RAPPORT), conceived to prioritise preventative interventions for people at risk of RA. PPI partners, including individuals at risk of rheumatoid arthritis (RA), RA patients, relatives and members of the public, provided feedback on key trial design issues through online meetings, a feedback form and emails. PPI processes employed during the one-year accelerator project were thereafter evaluated by PPI partners using an anonymous online feedback form with reference to National Institute of Health and Care Research (NIHR) UK standards for public involvement in research.

**Results:** Sixteen out of the 25-strong PPI partner panel completed an online feedback form (64%). Respondents perceived PPI processes positively in relation to all NIHR standard domains. Several key facilitators and challenges were identified, including the need for adequate PPI funding during pre-award phases of research, strategies for creating an inclusive environment, flexibility around levels of involvement, and challenges in achieving representatively diverse participation, and the importance of communicating transparent processes for role-assignment and time-reimbursement.

**Conclusions:** In general, RAPPORT was considered an example of PPI well done, and in line with UK standards for public involvement in research. Facilitators and challenges of relevance for the development of future translational and clinical trial funding applications are highlighted.

**PLAIN ENGLISH SUMMARY:** Patient and public involvement (PPI) in the development of funding applications to deliver clinical trials is desirable, but the PPI activities and processes involved at this early, “pre-award” stage are rarely reported. In the current paper we describe such activities during a 12-month project to develop a grant proposal for a substantive, international clinical trial. Three PPI partners were co-recipients of “seed funding” to conduct the 12-month Accelerator project, an additional 22 PPI partners being subsequently recruited to co-develop the funding application for the trial, entitled the “Rheumatoid Arthritis Prevention: catalysing PlatfORm Trial (RAPPORT).” PPI partners contributed through meetings, email discussions and the completion of feedback forms. The PPI processes used in the project were evaluated by 16 of the PPI partners using an anonymous online feedback form. The form asked about the areas covered by the UK Standards for Public Involvement.

PPI partners indicated that PPI in RAPPORT was done well in relation to all areas of the UK Standards. PPI partners felt they were heard, and their input valued, and that the communication was effective. Furthermore, they appreciated online format of the PPI activities, the flexible levels of involvement offered and the support from staff with expertise in both research and PPI. Some areas for potential improvement in future initiatives were also identified, which are discussed alongside challenges to co-development of projects during the “pre-award” stage in general, and the benefit of seed funding to support effective PPI.

## BACKGROUND

The involvement of patients and members of the general public at all stages of health research is increasingly recognised as an indication of good research practice (1, 2). The National Institute for Health and Care Research (NIHR), a major public funder of clinical trials in the UK, defines public involvement as “research being carried out ‘with’ or ‘by’ members of the public rather than ‘to’, ‘about’ or ‘for’ them. It is an active partnership between patients, carers and members of the public with researchers that influences and shapes research” (3). Evidence of active and meaningful Patient and Public Involvement (PPI) in the development of grant applications and in funded research projects is now a requirement for many public and charitable funding bodies, and specific UK standards for PPI have been created (4). Indeed, co-development of grant applications with PPI partners as co-applicants is encouraged with this in mind (5–7).

The value of such approaches is widely accepted (8–12), yet there are few detailed examples in the literature describing PPI happening in clinical trial design or translational research more generally (13). This is demonstrably the case in rheumatology research, for example, with PPI partners under-represented (14, 15) despite recommendations developed by The European Alliance of Associations for Rheumatology (EULAR) (16).

More particularly, although some guidance is available (e.g. (17)) very few detailed accounts of the *process* of involving PPI partners in the *development* of grant proposals, at the “pre-award” phase, or its evaluation, are available in the scientific literature to inform best practice. In the case of multi-centre and/or international projects, there may be specific PPI-related issues that need attention. For example, in cases where large and geographically diverse panels of PPI partners may be desirable, in-person attendance at meetings might not be possible for the PPI partners due to either (personal) travel restricts and/ or due to limited availability of pre-award funding for PPI costs associated with (inter)national travel. Whilst the use of remote, online PPI meetings may address some of these issues, this may present additional challenges, such as potential for digital exclusion or impaired relationship building.

We aimed to provide a detailed account and evaluation of PPI processes in a recent rheumatology endeavour to develop a substantive, collaborative funding application proposing an international platform trial of preventive interventions for people at risk of rheumatoid arthritis (RA): the Rheumatoid Arthritis Prevention: catalysing PlatfORm Trial (RAPPORT). In preparing this proposal, an international group of PPI partners was assembled, and several online PPI processes deployed, including online meetings and digital feedback forms to facilitate detailed PPI partner input and evaluate PPI activities. These preparatory activities were supported by “seed funding” in the form of a 12-month “accelerator” award, resourcing administrative elements and ensuring PPI partners could be offered reimbursement for their time and any expenses.

The main objectives of the current paper were to: 1) describe online PPI activities and processes in the development of the grant proposal for an international clinical trial; 2) describe feedback from PPI partners on these activities and processes; 3) reflect on alignment with NIHR national standards for PPI; and 4) describe key facilitators and challenges. The paper does not describe the trial protocol or the impact of PPI on the proposal as the application process is still ongoing.

## METHODS AND MATERIALS

### The RAPPORT project

A 12-month NIHR Efficacy and Mechanism Evaluation (EME) Accelerator Award (NIHR158397; September 2022-August 2023) was awarded to the Principal Investigator (AGP) to support development of the aforementioned substantive RAPPORT funding application to an MRC-NIHR EME Programme (18). Besides assembling an international network of investigators to design and deliver this trial, a stated objective of the accelerator project was to “convene and consult a RAPPORT Public Advisory Group to directly inform trial design, including a strategy for mapping the level of RA progression risk to lifestyle and/or pharmacological interventions.” (19) Approximately 10% of the Accelerator award was assigned to convene the group and to cover all PPI activities, PPI partner reimbursement and salary costs of dedicated PPI personnel across Newcastle and Birmingham Universities.

### Recruitment of an international PPI panel

The initial application for the accelerator award involved three PPI co-applicants affiliated with a pre-existing PPI group in the Rheumatology department of Newcastle upon Tyne Hospitals: the Patient and public Involvement and engagement in Musculoskeletal reSearch (PIMS) group (20). Each co-applicant brought lived experience as clinical trial participants in an RA prevention trial. Following the award of the accelerator funding, an international PPI panel (RAPPORT Public Advisory Group) was established (19).

Members of the international PPI panel were identified through their involvement as PPI partners in other pan-European projects addressing RA prevention (IMI-PREFER(21); IMI-RTCure (22); EuroTEAM (23)), as well as EULAR People with Arthritis and Rheumatism in Europe (PARE) networks and locally affiliated PPI groups (PIMS, the Birmingham Rheumatology Research Patient Partnership (R2P2) (24) and the Muscle Health PPI Group, Birmingham).

The PPI panel comprised 25 individuals and included 11 individuals with established RA of whom three reported prior lived experience of having been identified as at risk of RA, six individuals with arthralgia/joint problems but no diagnosis of RA, three first degree relatives of RA patients and two members of the public without joint problems or a diagnosis of RA. A further three PPI partners chose not to provide information about whether they had lived experience of being at an increased risk of developing RA or had a diagnosis of RA. The panel included PPI partners from the UK, Republic of Ireland and the Netherlands, and several partners had experience of being involved in (large-scale) international projects.

### Overview of PPI tasks and activities

After a discussion with PPI partners around their preferences for the meetings, it was decided that, in order to accommodate such a large and international group of PPI partners and ensure meetings/activities were widely accessible, all PPI activities would be conducted remotely. In addition to three online PPI meetings, PPI partners provided additional detailed feedback on key trial design issues (including recruitment procedure, informed consent procedure, intervention allocation and outcome prioritization) using an online form developed with input from the three accelerator award PPI co-applicants. The information gathered in this form remains the subject of ongoing trial development, and is not the focus of the present study, which instead addresses the PPI processes and approaches used.

The RAPPORT principle investigator (AGP), as well as a number of other researchers with PPI and scientific expertise were actively involved in all PPI meetings, development of feedback forms and other PPI activities, and were at hand to explain concepts further and provide guidance and assistance where needed.

Reasonable adjustments were made upon request to facilitate accessibility of PPI activities. For example, printed versions of electronic documents in large font were provided to some PPI partners. Meetings were recorded and transcribed so they could be shared after the event, and attendees were encouraged to enable any video cameras on their device when speaking, if possible, in an effort to optimise the quality of interaction. In addition, guides to the use of meeting platforms (MS Teams & Zoom) were provided in advance, to facilitate access.

PPI partners could choose which tasks and activities they got involved in for the duration of the one-year accelerator grant. *Table 1* gives a brief overview of PPI activities (see also the award report (19)). PPI partners were offered online vouchers for their time to participate in, and prepare for, PPI activities at a rate of £25 per hour.

**Table 1.** Overview of PPI tasks and activities: RAPPORT trial development.

|  |  |
| --- | --- |
| 1 | Developing the grant application for the accelerator award (n=3). |
| 2 | Attending an initial RAPPORT launch event (n=3). |
| 3 | Attending online meeting to kickstart the PPI panel (n=3) |
| 4 | Attending up to 2 online meetings during the lifetime of the accelerator grant (n=16-18). |
| 5 | Development of a novel online feedback form around key issues related to the trial design and grant application (n=2). |
| 6 | Giving feedback on the key trial design issues and stage 1 grant application via the online feedback form (n=22). |
| 7 | Involvement in the design of a study of patient treatment preferences to be |
|  | conducted during the trial (n=10). |
| 8 | Contributing to the development of a lay summary and preparation of PPI plans for the stage 1 NIHR grant application and beyond. (n=5) |
| 8 | Contributing to the preparation of a role description for PPI co-applicant roles in preparation for possible stage 2 grant submission. (n=5) |
| 9 | Completing an online feedback form to capture feedback on PPI activities and approaches undertaken during the accelerator grant. (n=16) |

### Evaluation of processes

On the suggestion of PPI partners during one of the online meetings, an anonymous online evaluation form was developed to gather feedback from PPI partners on their experiences (positive and negative) during their involvement with the RAPPORT accelerator project. The form was based on feedback questionnaires used in previous initiatives with a large PPI element (25, 26) and designed to address all areas of the NIHR national standards for PPI (4). These standards cover “Inclusive opportunities” (i.e., offering opportunities that are accessible to all relevant stakeholders; “Working together” (i.e. PPI partners and researchers collaborating in a way that values all contributions, and that builds and sustains mutually respectful and productive relationships); “Support and learning” (i.e., offering and promoting support and learning opportunities that build confidence and skills for public involvement in research); “Communications” (i.e., using plain language for well-timed and relevant communications regarding PPI); Impact (ensuring that PPI partners understand the impact of their involvement and that of PPI partners in general); and finally, “Governance” (i.e., involving PPI partners in research management, regulation, leadership and decision making) (4).

On the form, PPI partners were first asked to indicate whether they were a RA patient, a relative, someone with joint symptoms suspect of RA or a member of the public, how they were recruited to the PPI panel and what task(s) and activities they got involved in.

PPI partners subsequently indicated their level of agreement with 13 statements related to NIHR national standards using a 5-point rating scale (1 ‘strongly agree’ to 5 ‘strongly disagree’, as well as a ‘don’t know’ option). Example items included “I am satisfied with the amount of interaction I have had with the researchers” and “There was mutual respect between patient/public partners and researchers”.

This was followed by a section which assessed the use of the novel online form used to gather initial feedback from PPI partners on key trial design issues. This section consisted of five statements using the same 5-point rating scale and two open-ended questions: 1) “In what ways was the online feedback form an effective method of collecting feedback from patient/public partners about key aspects of the RAPPORT trial design?” and 2) “In what ways could the online feedback form on key aspects of the RAPPORT trial design have been improved?”.

The final section of the PPI form consisted of 11 open-ended questions related to the NIHR standards asking both what went well and what could be improved. For example, in relation to the ‘Inclusive opportunities’ standard we asked: “What went well, in relation to inclusivity and/or accessibility of the PPI panel?” and “What could have been improved, in relation to inclusivity and/or accessibility of the PPI panel?”. The final item provided an opportunity to add any further feedback. PPI partners were explicitly told that completion of these open-ended items was optional.

## RESULTS

A total of 16 PPI partners (64%), completed the online feedback form giving feedback on the PPI processes employed during the 1-year accelerator grant. All PPI partners indicated that they attended one or more online meetings. Ten also reported completing the online form gathering feedback on key trial design issues. Ten PPI partners were involved with the design of the patient preference study and 12 indicated they had been involved with developing the lay summary of the grant application. Responses to the rating scale items can be found in *Table 2* and are summarised below, organised by NIHR standard with supporting quotations from the open-ended questions.

**Table 2.**
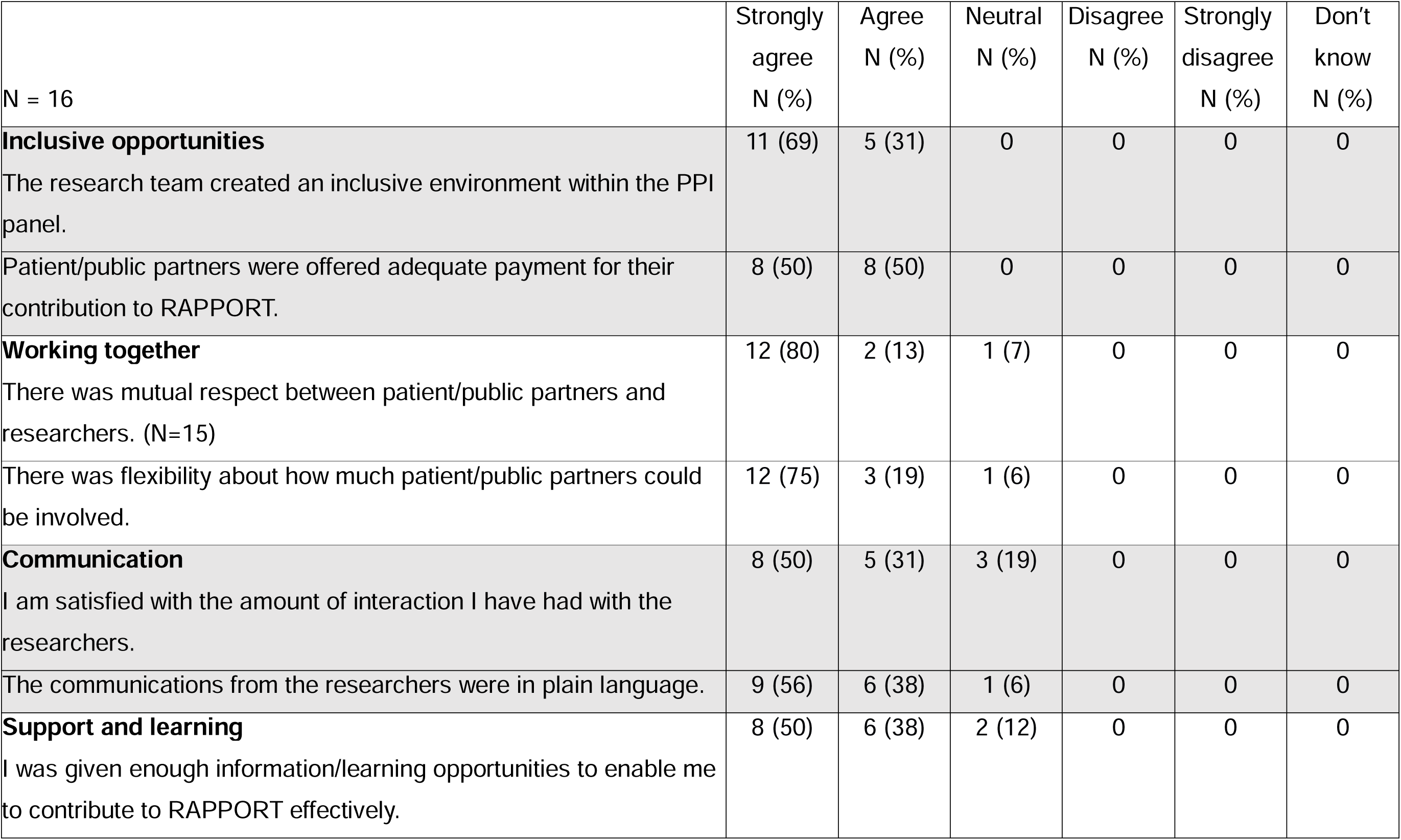
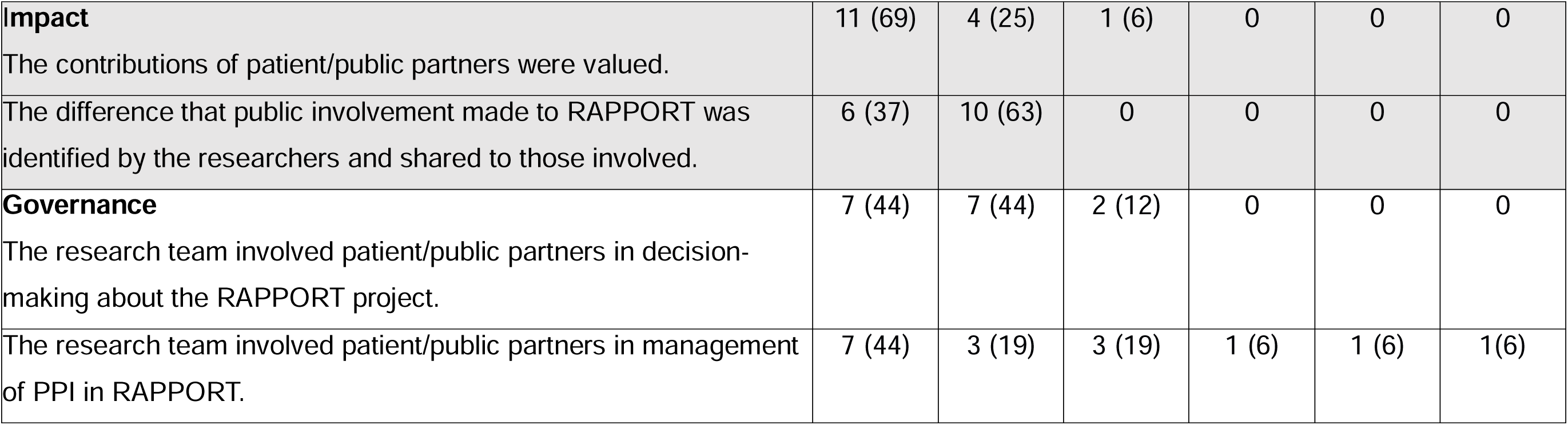
Agreement with statements related to the NIHR national standards.

All PPI partners felt that having been a PPI partner in RAPPORT had a positive impact on them, with ten PPI partners strongly agreeing, and six agreeing with the statement. Furthermore, almost all PPI partners would also recommend others to become involved in health research because of their experiences (strongly agreed n = 11; agreed n = 4) with one PPI partner indicating that they didn’t know whether they would or would not.

*“Happy with how it all went. Helped restore my faith in PPI. It’s actually probably been the best PPI I’ve experienced as a patient, and I’ve been involved for 10 years now! Another patient on the project said similar to me as well.”*

### Inclusive opportunities

All PPI partners strongly agreed (n = 11) or agreed (n = 5) that the research team created an inclusive environment within the PPI panel (see *Table 2*). As one PPI partner described:

> *“A relaxed and supportive environment where the leads gave the opportunity for all to contribute and actively made space and encouraged some of the quieter members of the group. A feeling of really being listened too and everyone’s contribution being valuable.”*

In addition, all PPI participants strongly agreed (n = 8) or agreed (n = 8) that payment offered for their PPI activities had been adequate (*Table 2*). However, one partner asked for a clearer payment strategy from the start of the project:

> *“I may be getting confused here with other projects, but I think in the beginning it wasn’t always clear if payment was being offered for an activity and how much, when and in what format this would be made. This has much improved during the course of the project.”*

Some respondents further indicated that it was difficult to say whether the PPI panel was thoroughly inclusive and diverse. For example, one PPI partner pointed out that no details were provided of the make-up of the PPI panel in terms of personal details such as sexual orientation, gender identity and ethnicity:

> *“I don’t know if we were fully inclusive as were not given details of the underrepresented groups in society – LGBTQ [lesbian, gay, bisexual, transgender, queer], different race and ethnicities, travellers [member of the community traditionally having a nomadic lifestyle] and BAME [Black, Asian and minority ethnic].”*

In general, having the meetings online and communication via email contributed to a feeling of inclusivity, with everyone being able to contribute regardless of location and ability to travel:

> *”It was very friendly and professional, and the ability to do the meetings online made it accessible for me, otherwise I would not have been able to do it. I found emails were answered quickly and nothing was too much trouble.”*

However, a perceived downside of having online meetings was a lack of ‘togetherness’. Online meetings were especially challenging for some when a number of participants did not join on camera:

> *“Found it difficult coping with contributors not on screen - just heard a voice. Found this very off-putting to the degree of a feeling of separation. Worked against togetherness.”*

It was also pointed out that people not being on camera made it more difficult to infer what was being said for those hard of hearing and a solution suggested was the use of online transcribing:

> *“Although researchers did repeat what had been said. Would have liked to have seen a transcript of what was being said in real-time on the screen. This would have helped.”*

Some respondents suggested offering additional face-to-face meetings, to increase the sense of belonging and allow for networking:

> *“Maybe have one face-to-face meeting so everyone can network.”*

Others suggested providing more choice in meeting time and/or providing evening slots to increase inclusivity to those working:

> *“Finding out from PPI partners what the best time would be to schedule meetings/sessions. Identifying a range of 3 dates/times to allow the PPI partner to indicate which is convenient and then going with the one that includes as many as possible.”*

### Working together

Most respondents (87%) indicated that there was mutual respect between PPI partners and researchers (n = 12 strongly agreed; n = 2 agreed). Similarly, almost all (94%) felt that there was flexibility around how much PPI partners could be involved (n = 12 strongly agreed; n = 3 agreed; see also *Table 2*):

> *“Respectful attitude of researchers and clearly communicated, timely feedback. Good notice of events/future input and reassurance that input could vary given one’s personal time and circumstances. Clear emails that detailed their plans and needs with adequate notice, not needy requests late in the day, that I have experienced with other projects. I think the researchers showed genuine interest and motivation to improve the quality of PPI in all research, not just their own. Prompt feedback of results/findings.”*

There were some suggestions for improvements or future work as well, for example some indicated that either more meetings or smaller group meetings might be desirable, in this context the desire for a role description was also mentioned:

> *“A role description and we could have been split into different groups to have discussions to bring more richness from the lived experience to the project.”*

### Communications

A large proportion of PPI partners (81%) were satisfied with the amount of interaction they had with the researchers (n = 8 strongly agreed; 5 = agreed) and most (94%) agreed that the communications were in plain language (n = 9 strongly agreed; 6 = agreed; see also *Table 2*).

> *“Very happy. I felt the communications were clear, not too frequent or overwhelming. Also, given enough time to review info (especially before meetings). Am quite used to bad practice of disorganised research teams sending tonnes of info to read at the 11th hour.”*

However, some respondents would have liked more (detailed) emails and others felt that researchers should have checked which communication means were preferred:

> *“You may want to check to see what is the preferred communication and how to be contacted.”*

### Support and learning

Most PPI partners who completed the PPI feedback form (87%) felt that they were given enough information/learning opportunities to enable them to contribute to RAPPORT effectively (n = 8 strongly agreed; n = 6 agreed; see *Table 2*). Some felt that what was offered was enough, others that they simply did not need additional training:

> *“I believe my common sense is the best attribute I bring to PPIE [patient and public involvement and engagement] and other than procedural guidance I do not want to be swamped with too much of the science.”*

Others indicated that offering additional information about training might increase PPI input from individuals who might normally not come forward:

> *“I like the way PRPs [patient research partners] could opt to help at different levels. Indicating which of these levels might come with some extra information/training might be helpful in recruiting participation from individuals who may question their ability to contribute depending on their direct/indirect experience of the disease, knowledge and experience of research.”*

### Impact

Most PPI partners (94%) felt that the contributions of patient/public partners were valued (n= 11 strongly agreed; n = 4 agreed). All partners reported that the difference that public involvement made to RAPPORT was identified by the researchers and shared to those involved (n = 6 strongly agreed; n = 10 agreed; see also *Table 2*).

The respondents further highlighted the ways they felt the researchers shared impact:

> *“They gave examples of when decisions had been made with specific input from patient partners.”*

> *“I think this was done well and feedback both via meetings and email correspondence. This included both when things had gone well or not as hoped, this felt very honest and open.”*

One PPI partner felt impact was perhaps not communicated effectively, although always acknowledged:

> *“I don’t think this was the case. However, we were always thanked and acknowledged our contributions.”*

Some suggestions on how to measure PPI impact were also made, for example:

> *“Measure the impact through infographic - time, number of people involved what achieved and any other outputs/outcomes.”*

### Governance

Most PPI partners (87%) felt that the research team involved patient/public partners in decision-making about the RAPPORT project (n = 7 strongly agreed; n = 7 agreed; see *Table 2*) and over half (62%) indicated that the research team involved patient/public partners in management of PPI in RAPPORT (n = 7 strongly agreed; n = 3 agreed). However, three partners were neutral on the subject and two disagreed or strongly disagreed with the statement (see also *Table 2*).

PPI partners described how they felt their views influenced the decisions ultimately made by the research team and made a difference:

> *“The views that were offered by PRPs were clearly utilised and helped shape decisions that were made by the research team.”*

> *“I believe we made a difference, that our voices were heard, and it helped to shape the project.”*

One PPI partner suggested there was scope for increased transparency in the decision-making process:

> *“Possibly a discussion about how decisions were made/agreed. How PPI decisions/researchers’ decisions could be balanced if contrary feelings. I.e., would PPI and researcher ideas have equal standing.”*

### Use of an online form to gather feedback on key issues of the research design

As part of one of the PPI activities an online form was used to gather feedback on various key issues of the trial design. Twelve of the 16 respondents (75%) who completed the PPI evaluation form felt that this key issue feedback form was ‘an effective way of getting input on key aspects of the research design’ (n = 5 strongly agree; n = 7 agree; see also *Table 3*).

**Table 3.**
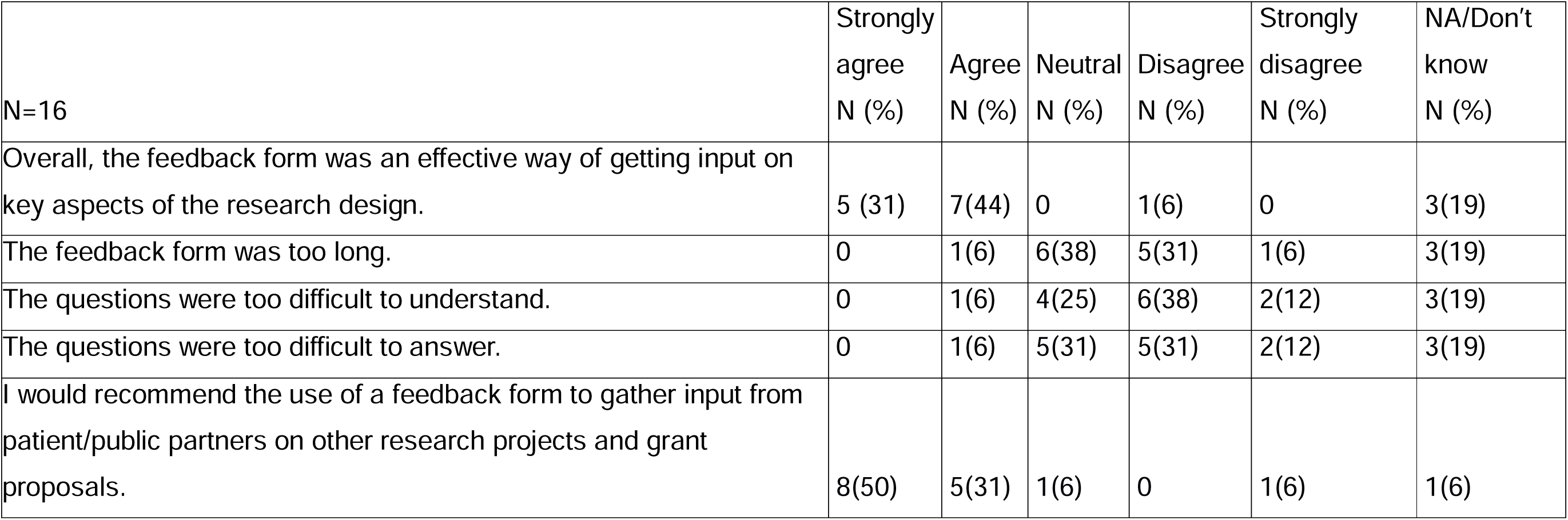
Agreement with statements related to use of feedback form to get input on research design:

|  | Strongly<br>agree<br>N (%) | Agree<br>N (%) | Neutral<br>N (%) | Disagree<br>N (%) | Strongly<br>disagree<br>N (%) | NA/Don't<br>know<br>N (%) |
| --- | --- | --- | --- | --- | --- | --- |
| N=16 |  |  |  |  |  |  |
| Overall, the feedback form was an effective way of getting input on key aspects of the research design. | 5 (31) | 7(44) | 0 | 1(6) | 0 | 3(19) |
| The feedback form was too long. | 0 | 1(6) | 6(38) | 5(31) | 1(6) | 3(19) |
| The questions were too difficult to understand. | 0 | 1(6) | 4(25) | 6(38) | 2(12) | 3(19) |
| The questions were too difficult to answer. | 0 | 1(6) | 5(31) | 5(31) | 2(12) | 3(19) |
| I would recommend the use of a feedback form to gather input from patient/public partners on other research projects and grant proposals. | 8(50) | 5(31) | 1(6) | 0 | 1(6) | 1(6) |

Only one respondent indicated that the key issue feedback form was too long, the questions were difficult to understand and difficult to answer. Three partners indicated that they could not answer these questions and around a quarter was neutral on the subject of understanding and difficult to answer questions (n =4, n=5 respectively).Thirteen partners (81%) recommended the use of a similar feedback form to gather input from PPI partners on key issues in other research projects and grant proposals (n = 8 strongly agree; n = 5 agree; see also *Table 3*).

In their written feedback, PPI partners further suggested that the online feedback form was accessible, and allowed more individuals to provide feedback on the trial design; although it could be improved by being more concise, having additional information or being available in other languages:

> *“Accessible by all those who can gain access to computer. Allows relative freedom when completed, therefore flexible to patient/public needs and availability. Allows independent viewpoints to be heard, rather than be led by others with louder voices/opinions. Allows those who find it hard in person or with audience to contribute. Gives anonymity. May allow those who’s [whose] first language [is] not English to have time to seek help from others to understand the questions and give feedback.”1*

## DISCUSSION

The main objectives of the current paper were to describe and evaluate online PPI activities and processes in the development of the grant proposal for an international clinical trial and to reflect on alignment with NIHR national standards for PPI.

The PPI processes within this project were perceived positively in relation to all domains of the NIHR standards and more than 80% of respondents agreed or strongly agreed with the positive statement in most cases, except for one rating relating to governance where 62% of respondents agreed or strongly agreed with the positive statement.

The PPI panel was considered to be an inclusive environment where PPI partners were listened to and felt respected by the researchers and PPI input was incorporated into the grant proposal. The general consensus was that the project was an example of PPI done well. However, some key facilitators and challenges were highlighted during the project, which are discussed below.

### Key facilitators

#### Pre-award funding for PPI

In the accelerator award described in this manuscript, adequate funding was available to offer reimbursement to a large panel of PPI partners for their time and expenses. This allowed us to set up a varied panel of PPI partners (as recommended by EULAR (27)) and provided flexibility for PPI partners in relation to their level of involvement in PPI activities.

Involving PPI partners from the concept stage of a proposal allows them to evaluate the relevance/direction of a proposed study rather than being confronted by a more mature proposal as a *fait accompli*, beyond the point at which PPI input is likely to have a meaningful impact on priority-setting. Involvement only at later, post-award stages may thereby impede fruitful interaction between researchers and PPI partners whose vision for the research is not aligned, reducing overall levels of involvement.

PPI partners appreciated the payment offered for PPI activities as a recognition of their time and effort as well as the dedicated PPI facilitators for the project. Without the accelerator funding, this would not have been possible. However, although there is a welcome emerging trend for large, collaborative funding opportunities to be preceded by seed funding from, for example, the NIHR(18), it is still unusual for funding of this nature to be available during the pre-award stage of a project. Amongst smaller funders especially, such funding is unlikely to become available due to financial and logistical constraints.

#### PPI supported by staff with both scientific and PPI expertise

For the current project funds were deployed to employ staff with both research and PPI expertise, to coordinate PPI activities. The extent and quality of PPI that occurred in this funding application would not have been possible without appropriate resource to support staff with expertise in PPI coordination and communication, and with access to established networks of PPI partners with specialist interest in RA prevention research. Access to existing PPI groups or networks with PPI partners who have some experience with PPI from previous projects and/or have received relevant training. allows for a matching of PPI partners skills and areas of interest with specific projects.

Furthermore, the Principal Investigator was involved in most of the PPI activities providing further scientific expertise. Although not directly captured in the feedback form, PPI co-authors highlighted that this active engagement was perceived to be very beneficial. The need for senior researchers with both scientific and PPI expertise to facilitate successful PPI has also been highlighted in other international projects (13).

#### Online platforms

The use of online platforms for meetings (with appropriate guidance), and email and online forms for further feedback facilitated the inclusion of a large, international panel of PPI partners. Conducting most PPI activities online reduces the need for travel and associated costs. It further allows everyone to contribute to the discussions in a variety of ways, for example via “chat” functions that circumvent the need to speak up during the meetings for those individuals who are less comfortable doing so, or via email after the meeting, perhaps in reaction to transcripts. This does, however, present risk of digital exclusion, as discussed in more detail below as a key challenge.

Using an online form for PPI partners to give detailed feedback on key considerations in the grant proposal proved successful in the current project, and this relatively novel approach has gained traction during and since the COVID-19 pandemic (13). PPI partners particularly valued it as a means of increasing inclusivity, since everyone had an opportunity to provide feedback both through brief rating scales and through more detailed answers to open-ended questions at their own pace. However, a close eye needs to be kept on both the length of any such form as well as its complexity as exemplified by some of the more negative ratings regarding the form by some of the PPI partners.

#### Flexible levels of involvement

In the current project, PPI partners particularly appreciated flexibility around levels of involvement and the ability to choose what activities to get involved in and to what extent according to their availability, interest and skillsets. Having a relatively large panel of PPI partners avoided feelings of obligation to commit to all PPI activities and allowed for varied levels of commitment (robust to drop-outs), maximizing overall involvement.

### Key challenges

#### Enrolment of a representatively diverse panel

When asked about diversity in the evaluation form, some PPI partners indicated that they felt they could not really assess the diversity of the current PPI panel as the information was not readily available, but that there is need for diversity and the inclusion of underrepresented groups in PPI panels in terms of ethnicity, sex, sexual orientation, and gender identity. This is a key consideration to ensure that research objectives and methods are inclusive and appropriate for diverse populations. Indeed, this information was not captured for the PPI panel recruited during the accelerator grant and, although we recruited our PPI partners through a variety of channels and locations, we cannot appraise objective data to determine whether this resulted in greater diversity of our PPI panel.

As noted by others, PPI partners in the UK tend to be white, female retirees (**28**). To address this limitation in future projects it would first be necessary to capture protected characteristics data on prospective PPI panel members at the point of enrolment whilst ensuring this is done in a way that complies with general data protection regulation (GDPR) legislation. Transparency about the reasons for gathering this data, allowing opt out and ensuring appropriate data protection is necessary to support disclosure and collection of data with which to monitor the diversity of PPI panels in both pre-award and post-award phases of research. Time constraints, in addition to funding constraints, are typically key barriers to implementing this more comprehensive approach to equality, diversity and inclusion when convening PPI partners during the pre-award stage of funding application development.

#### Digital exclusion mitigation

Online meetings were often seen as convenient, and a positive means to enhance inclusivity. However, reliance on online meetings, feedback forms and email communications does mean that those with no access to computers, problems using computers or who are simply not comfortable with digital technology could inadvertently be excluded/ disadvantaged.

During online meetings participants can be encouraged to have their cameras on (unless this is not possible for personal, medical or technical reasons). Providing information on how to use automated captioning during the meeting and providing recordings and transcripts of the meetings after the meeting can further enhance accessibility.

However, the value of in person and/or smaller group meetings shouldn’t be overlooked in this context. There were suggestions for (additional) face-to-face meetings, meetings with smaller groups and/or the use of breakout rooms in the online meetings to allow PPI partners to network and increase engagement and productivity.

Having at least an initial face-to-face or hybrid meeting at the start of a project, as well as providing the option of telephone consultations and/or comments on paper copies of documents, might counteract some of the disadvantages and enhance inclusivity and accessibility. However, this would be cost- and time-intensive for both the researchers and PPI partner(s) in question unless substantial grant development funding is available. In the absence of such funding, early discussion with PPI partners to identify and address their communication needs and preferences is essential.

#### Clear communication and transparency

PPI structure and role in governance was not perceived as entirely clear to the whole group of PPI partners in the current project from the outset. For example, not every participant accepting an invitation to join the wider PPI panel was aware that three of their counterparts were co-applicants for the accelerator “seed funding” that underpinned the project. Neither was co-development of a clear PPI role description and its implementation formally undertaken for the immediate benefit of the panel during this pre-award period – although its formulation for purposes of the substantive RAPPORT application was an output of the accelerator work.

Furthermore, whilst participants generally considered the compensation offered for their time commitment to be adequate, and the payment structure became apparent after the start of the project, these elements were not explicitly communicated in the initial invitation. Ideally, such details should be made clear and transparent from the earliest possible stage at which a prospective PPI partner becomes involved in a project. It should be remembered, however, that during these early stages of development, formal governance frameworks for a given research project are typically in evolution as the research and delivery team itself convenes, and an iterative element to PPI considerations is, arguably, inevitable.

With respect to time reimbursement, it should nonetheless normally be possible to indicate arrangements promptly and, particularly where there are no funds to pay for pre-award PPI activities (as is often the case), this should be communicated so prospective PPI partners can make an informed decision as to whether they would like to be involved. Co-development of PPI role descriptions, setting out relevant PPI processes for a particular project and a summary of PPI activities and tasks, should be one of the first activities/discussions to be undertaken with a newly formed PPI panel.

## CONCLUSION

The general consensus amongst a PPI panel convened for the pre-award development phase of the substantive, international clinical trial proposal described in this paper was that PPI aligned with best practice guidelines. This was facilitated by the availability of dedicated pre-award seed funding, which is uncommon in research proposal development. Challenges of implementing PPI for a large, international panel of PPI partners related to reliance on online interactions, and the need to mitigate for potential digital exclusion.

## Data Availability

The data that support the findings of this study are available on request from the corresponding author. The data are not publicly available due to privacy restrictions.

## LIST OF ABBREVIATIONS

BAME: Black, Asian and minority ethnic
BRC: NIHR Birmingham Biomedical Research Centre
COVID-19: Coronavirus disease
EME: Efficacy and Mechanism Evaluation
EULAR: The European Alliance of Associations for Rheumatology
GDPR: general data protection regulation
LGBTQ: lesbian, gay, bisexual, transgender, queer
NIHR: The National Institute for Health and Care Research
PIMS: Patient and public Involvement and engagement in Musculoskeletal reSearch
PPI: Patient and public involvement
PPIE: Patient and public involvement and engagement
PRP: patient research partner
R2P2: Birmingham Rheumatology Research Patient Partnership
RA: rheumatoid arthritis
RAPPORT: Rheumatoid Arthritis Prevention: catalysing PlatfORm Trial
REC: research ethics committee
UK: United Kingdom

## DECLARATIONS

### Ethics approval and consent to participate

In this paper, we report on the evaluation of PPI processes used during the development of a NIHR Phase 1 grant application for a clinical trial. PPI partners were involved in all PPI activities, co-developed an online form to gather feedback on key issues of the trial and suggested the evaluation of PPI processes using an online form. All respondents were PPI partners, were aware of the purpose of the evaluation, and how the findings would be used. This was a PPI activity, for which neither research ethics committee (REC) nor formal informed consent procedure is required according to Health Research Authority guidelines in line with the UK Policy Framework for Health and Social Care Research (29). All methods were carried out in accordance with relevant guidelines and regulations, including the UK Standards for Patient and Public Involvement in Research (4).

### Consent for publication

Not applicable

### Author contributions

GS: Conceptualization; Methodology; Writing first draft manuscript; Writing further drafts; Formal statistical analysis of evaluation exercise; Qualitative analysis of evaluation exercise. MF: Conceptualization; Methodology; Extensive review and editing first draft manuscript; Review of further drafts; Agreement final draft manuscript; Qualitative analysis of evaluation exercise. AGP: Conceptualization; Methodology; Extensive review and editing first draft manuscript; Review of further drafts; Agreement final draft manuscript. All other authors: Conceptualization; Methodology; Manuscript review and editing; Agreement final draft manuscript.

## Acknowledgements

We would like to thank all PPI partners who have been part of the RAPPORT PPI panel, both PPI co-authors and those who choose not to be directly involved with this publication but have extensively contributed to the PPI activities. In particular, we would like to thank Barbara Usherwood and Peter Armstrong who together with Helen Jones were PPI co-applicants for the accelerator-grant application funding this work.

## REFERENCES

1. Blackburn S, McLachlan S, Jowett S, Kinghorn P, Gill P, Higginbottom A, et al. The extent, quality and impact of patient and public involvement in primary care research: a mixed methods study. Research Involvement and Engagement. 2018;4(1):16.

2. Aiyegbusi OL, McMullan C, Hughes SE, Turner GM, Subramanian A, Hotham R, et al. Considerations for patient and public involvement and engagement in health research. Nature Medicine. 2023;29(8):1922–9.

3. NIHR. Briefing notes for researchers - public involvement in NHS, health and social care research 2021 [updated April 2021. 1:[Available from: https://www.nihr.ac.uk/documents/briefing-notes-for-researchers-public-involvement-in-nhs-health-and-social-care-research/27371.

4. NIHR. UK standards for public involvement [Available from: https://sites.google.com/nihr.ac.uk/pi-standards/home.

5. de Wit M, Teunissen T, van Houtum L, Weide M. Development of a standard form for assessing research grant applications from the perspective of patients. Research Involvement and Engagement. 2018;4(1):27.

6. Ní Shé É, Morton S, Lambert V, Ní Cheallaigh C, Lacey V, Dunn E, et al. Clarifying the mechanisms and resources that enable the reciprocal involvement of seldom heard groups in health and social care research: A collaborative rapid realist review process. Health Expectations. 2019;22(3):298–306.

7. Ní Shé É, Cassidy J, Davies C, De Brún A, Donnelly S, Dorris E, et al. Minding the gap: identifying values to enable public and patient involvement at the pre-commencement stage of research projects. Research Involvement and Engagement. 2020;6(1):46.

8. Birch R, Simons G, Wähämaa H, McGrath CM, Johansson EC, Skingle D, et al. Development and formative evaluation of patient research partner involvement in a multi-disciplinary European translational research project. Research Involvement and Engagement. 2020;6(1):6.

9. Simons G, Birch R, Stocks J, Insch E, Rijckborst R, Neag G, et al. The student patient alliance: development and formative evaluation of an initiative to support collaborations between patient and public involvement partners and doctoral students. BMC Rheumatology. 2023;7(1):36.

10. Smith MY, Janssens R, Jimenez-Moreno AC, Cleemput I, Muller M, Oliveri S, et al. Patients as research partners in preference studies: learnings from IMI-PREFER. Research Involvement and Engagement. 2023;9(1):1–14.

11. Biggane AM, Olsen M, Williamson PR. PPI in research: a reflection from early stage researchers. Research Involvement and Engagement. 2019;5(1):35.

12. Hewlett S, de Wit M, Richards P, Quest E, Hughes R, Heiberg T, et al. Patients and professionals as research partners: Challenges, practicalities, and benefits. Arthritis Care & Research. 2006;55(4):676–80.

13. de Souza S, Johansson EC, Karlfeldt S, Raza K, Williams R. Patient and public involvement in an international rheumatology translational research project: an evaluation. BMC Rheumatology. 2022;6(1):83.

14. Elhai M, Benavent D, Aouad K, Studenic P, Bertheussen H, Primdahl J, et al. Involving patients as research partners in research in rheumatology: a literature review in 2023. RMD Open. 2023;9(4):e003566.

15. Wang H, Stewart S, Darlow B, Horgan B, Hosie G, Clark J, et al. Patient research partner involvement in rheumatology clinical trials: analysis of journal articles 2016-2020. Ann Rheum Dis. 2021;80(8):1095–6.

16. de Wit M, Aouad K, Elhai M, Benavent D, Bertheussen H, Blackburn S, et al. EULAR recommendations for the involvement of patient research partners in rheumatology research: 2023 update. Annals of the Rheumatic Diseases. 2024:ard-2024-225566.

17. Hoddinott P, Pollock A, O’Cathain A, Boyer I, Taylor J, MacDonald C, et al. How to incorporate patient and public perspectives into the design and conduct of research. F1000Res. 2018;7:752.

18. NIHR. Efficacy and Mechanism Evaluation 2023 [Available from: https://www.nihr.ac.uk/explore-nihr/funding-programmes/efficacy-and-mechanism-evaluation.htm.

19. Bardgett M, Falahee M, Simons G, Isaacs JD, Ouma L, Wason J, et al. Rheumatoid Arthritis Prevention: catalysing PlatfORm Trial delivery (RAPPORT). 2024.

20. NIHR Newcastle Biomedical Research Centre. Patient and public Involvement and engagement in Musculoskeletal reSearch (PIMS) [Available from: https://www.newcastlebrc.nihr.ac.uk/patients-carers-public/pims/.

21. European Commission. Patient Preferences in benefit risk assessments during the drug life cycle - Sofia ref.: 115966 2016 [updated 16/12/2022; cited 2024. Available from: https://cordis.europa.eu/project/id/115966.

22. European Commission. Rheuma Tolerance for Cure 2017 [updated 18/12/2023. Available from: https://cordis.europa.eu/project/id/777357.

23. European Commission. Towards Early diagnosis and biomarker validation in Arthritis Management 2012 [updated 10/09/2017; cited 2024. Available from: https://cordis.europa.eu/project/id/305549.

24. University of Birmingham. Birmingham Rheumatology Research Patient Partnership (R2P2) [Available from: https://www.birmingham.ac.uk/research/inflammation-ageing/research/r2p2/index.aspx.

25. Simons G, Birch R, Stocks J, Insch E, Rijckborst R, Neag G, et al. The Student Patient Alliance: Development and formative evaluation of an initiative to support collaborations between patient and public involvement contributors and doctoral students. medRxiv; 2023.

26. Birch R, Simons G, Wähämaa H, McGrath CM, Johansson EC, Skingle D, et al. Development and formative evaluation of patient research partner involvement in a multi-disciplinary European translational research project. Res Involv Engagem. 2020;6:6.

27. de Wit M, Berlo SE, Aanerud GJ, Aletaha D, Bijlsma JW, Croucher L, et al. European League Against Rheumatism recommendations for the inclusion of patient representatives in scientific projects. Annals of the Rheumatic Diseases. 2011;70(5):722–6.

28. Keane A, Islam S, Parsons S, Verma A, Farragher T, Forde D, et al. Understanding who is and isn’t involved and engaged in health research: capturing and analysing demographic data to diversify patient and public involvement and engagement. Research Involvement and Engagement. 2023;9(1):30.

29. NHS Health Research Authority. What do I need to do? 2020 [Available from: https://www.hra.nhs.uk/planning-and-improving-research/best-practice/public-involvement/what-do-i-need-do/#~:text=Do%20I%20need%20HRA%20ethical,people%20involved%20are%20NHS%20patients.

